# Clinical Response to fMRI-guided Compared to Non-Image Guided rTMS in Depression and PTSD: A Randomized Trial

**DOI:** 10.1101/2024.07.29.24311191

**Authors:** Desmond J. Oathes, Almaris Figueroa Gonzalez, Julie Grier, Camille Blaine, Sarai D. Garcia, Kristin A. Linn

## Abstract

**Background:** Image-guided brain stimulation is hypothesized to enhance clinical outcomes but head-to-head comparisons favoring image-guidance are so far lacking.

**Methods:** PTSD/MDD patients were randomized (N=51) to a two-condition sequence in a two period cross-over design. For the first condition, patients were randomized to 10-session rTMS treatment to either a subgenual cingulate (sgACC) functional connectivity cortical target (fMRI-guided) or standard scalp-based target. Additionally, patients were randomized to either watch a nature video or perform a demanding cognitive task with rTMS administration. Patients crossed over to the two conditions not received in period one. rTMS was delivered in an intermittent theta burst (iTBS) pattern with 2400 pulses per session. Among N=49 patients analyzed, 60% identified as female and average age was 34.

**Results:** Compared with the scalp-based target, fMRI-guided rTMS was superior in improving depression symptoms (*F*(1,43.92)=5.933, *p*=0.019) as well as PTSD hyperarousal (*F*(1,40.78)=5.076, *p*=0.030). The median level of symptom change for fMRI-guided targets exceeded 60% improvement across both scales. Symptom improvements at 6-mo follow-up were durable and both favored fMRI-guidance. For patients reporting symptoms at this timepoint, depression improved by 70% (N13); the PCL improved by 69% with Hyperarousal (N14) and Avoidance (N12) subscales improving by 78% and 79%, respectively, for the fMRI-guided target.

**Conclusions:** We demonstrated preliminary evidence for the clinical superiority of a new fMRI-guided target which should be followed up with larger comparative effectiveness studies that include imaging and clinical outcomes.

## Introduction

Neuroimaging-guided brain stimulation is seen as state-of-the-art for personalized medicine across a variety of indications in neuropsychiatry. However, large clinical trials of transcranial magnetic stimulation (TMS) have so far failed to demonstrate neuroimaging-defined target superiority compared to less anatomically precise scalp-based targets for treating depression.^1,2^ A recently FDA-approved protocol that utilized a functional MRI (fMRI)-guided target based on subgenual anterior cingulate (sgACC) connectivity demonstrated clinical effects substantially stronger than those reported in prior scalp-based target studies.^3,4^ That protocol also delivered repetitive TMS (rTMS) in a substantially different dose and timing, leaving open the question of whether the fMRI-guided targeting was essential to drive the impressive clinical effects. The fMRI-guided target in the present study was also the sgACC, but it was defined using positive functional connectivity (FC) connections rather than negatively correlated connections explored in most prior studies, which we chose in light of recent interleaved TMS/fMRI evidence for superior sgACC engagement in patients using this target.^5^

The sgACC has been shown to be consistently hyperactive in major depression disorder (MDD)^6^ as well as in post-traumatic stress disorder (PTSD),^7^ likely due to diagnostic and symptom overlaps between the two conditions.^8,9^ Clinical improvements following rTMS treatment are associated with sgACC connectivity changes on both depression and PTSD symptom scales.^10-12^ Recent studies by our group have shown that resting fMRI-guided TMS can effectively engage the sgACC in both patients and healthy controls.^13,14^ Considering this evidence, coupled with the fact that MDD and PTSD are the most frequent psychiatric conditions resulting from trauma, we included patients with trauma-induced (or exacerbated) MDD as well as PTSD in our study.

In this study, we contrasted the effects of an fMRI-based target with a scalp-based estimate of the dorsolateral prefrontal cortex (DLPFC) 6 cm anterior to the motor cortex^15^ on clinical depression and PTSD symptoms. The original targeting scheme centers the TMS coil 5 cm anterior to the motor cortex along a parasagittal line.^16^ Although this approach is simple to perform, recent studies have found significant discrepancy between the target identified by the 5 cm rule and targets guided by stereotaxic methods. A modified 6 cm rule for effective DLPFC stimulation was subsequently proposed to increase the likelihood of targeting the DLPFC in most participants.^17,18^ Although we hypothesized clinical benefits of the fMRI-guided targets over the scalp-based targets, we additionally sought to explore factors that might further improve symptoms across both targeting strategies. Specifically, we examined the effects of brain-state dependence (concurrent tasks) on clinical outcomes.

## Materials and methods

### Overview

Following a phone screen, diagnostic interview, symptom assessment, and a baseline MRI session to determine fMRI-guided targets, patients were randomly assigned to a sequence of two stimulation sites and, separately, a sequence of two experimental manipulations (tasks) in a two-period crossover design. The N-back task was chosen as one manipulation to induce a higher cognitive demand, higher arousal, and stress brain state; the other task was a video designed for patients to watch to induce a calmer, lower arousal brain state. Across the two periods, each of which involved a two-week rTMS session, participants were initially treated with one of two stimulation sites and one of two brain states. Then, both conditions were switched within-subject for the 2^nd^ rTMS session.

### Patients

Fifty-one unmedicated patient participants (see Supplement for demographics, baseline symptoms, diagnoses, and antidepressant resistance details) who met DSM-5 Diagnosis of MDD and/or PTSD were recruited and randomized (see Supplement for CONSORT diagram and exclusion criteria) through the University of Pennsylvania, Veterans Administration Medical Center, and surrounding community.

All participants gave consent for the experiment according to the Declaration of Helsinki, and the study protocol was approved by the institutional review board of University of Pennsylvania. The study was registered with ClinicalTrials.gov as NCT03114891 with a start date of 4/20/17 and a study completion date of 12/01/21.

### MRI acquisition and processing

For each participant, resting state fMRI and high resolution anatomical (T1) scans were acquired identical to our prior work^14^ for neuronavigated TMS targeting (see Supplement).

### TMS treatment

TMS was delivered using an MRI compatible Magventure B65 air-cooled TMS coil triggered by a Magpro X100 stimulator (Magventure; Farum, Denmark). The iTBS protocol was set using normal biphasic bursts of 3 pulses at 50-Hz (20ms inter-pulse interval) at a repetition rate of 5 pulses (triplets) per second (5 Hz), and an 8s inter-train interval for 40 trains (1,200 pulses) following a break for the task (∼10 min; see Supplement Methods) followed by a 2^nd^ round of the same protocol for a total of 2,400 pulses per session delivered at 70% resting motor threshold (see Supplement; ∼12.5 min of rTMS delivery).

The stimulation site was projected onto the structural image for each participant, and neuronavigation software (Brainsight; Rogue Research, Montreal, Quebec, Canada) was employed to ensure TMS coil position on the stimulation target in real time.

### TMS targets

Identification of the DLPFC target was achieved through connectivity-based targeting (fMRI-guided) or following the 6 cm rule (6cm). The fMRI-guided target was localized by a peak correlation cluster seeding the sgACC in the left DLPFC cortical surface region. The peak was not a single voxel; only sites with homogenous zones of positive valence FC values within ∼1cm of the peak were selected. When choosing between FC peak blobs in DLPFC, larger representations closer to the brain surface in zones closer to the scalp surface were selected. The 6 cm target was determined at a site 6 cm anterior to the M1 along the parasagittal line. Each participant had treatment to the fMRI-guided and 6 cm target with the sequence randomly assigned to participants as part of the crossover design.

### Clinical outcome scales

MDD and PTSD symptoms were assessed by the Patient Health Questionnaire (PHQ9)^19^ and the PTSD Checklist for DSM-5 (PCL-5),^20^ respectively. These scales were measured at baseline, every treatment visit, and at one- and two-week follow-ups after the treatment was completed for each treatment period. Clinical follow-up measures were also collected monthly up to 6-months after completion of the 2^nd^ intervention to assess the longevity of clinical improvement. Fourteen patients were missing a PHQ9 score from the baseline visit, so the PHQ9 score obtained at the screening visit was carried forward and served as a baseline for those patients.

### Randomization and blinding

Randomization of the target and the task sequence jointly across the two factors was conducted before the first treatment period started using a random number generator (Excel ‘rand’). The sequence of sites and tasks was not blind to the clinician or patient. The clinician determined the 6cm target and so could tell which of the two sites was scalp-derived. However, the clinician was not included in scientific project discussions and therefore was not aware of how the additional (non-6cm) site was generated nor its evidence basis. Analysts were not blinded to target or task assignment.

### Statistical analysis

On average, subjects experienced large reductions in symptoms throughout the first 2-week treatment period that did not return to pre-treatment symptom levels before the start of the 2^nd^ 2-week treatment period. Due to this observed carryover of symptom reductions (see Supplemental Results), we focused our analysis solely on data from the 1^st^ treatment period including clinical scores collected at baseline; days 2, 4, 6, 8, and 10; and the period 1 one-week follow up visit. At each timepoint, all available patient observations were used to plot symptom changes from baseline, i.e., complete cases by timepoint. We report rates of response by stimulation site and by task (cognitive task versus nature video) throughout the 1^st^ treatment period below.

Longitudinal trajectories of clinical outcome scales were modelled as change from baseline using linear mixed models. In all models, we controlled for the baseline clinical score as a fixed effect. Observation times between measures varied slightly (i.e., when treatment visits occurred; see Supplement). Time in days from baseline was included as a linear fixed effect in all models, utilizing the actual timing each patient completed their treatment visits, i.e., when the clinical outcomes were collected. All models included a random subject-level intercept. PHQ9, PCL-5, and PCL sub-scores were modelled separately.

For each clinical outcome, if the site-by-task interaction was not significant, we dropped the interaction term, refit the model, and tested for main effects of site and task. If the site-by-task interaction was significant, we tested for a significant effect of site within levels of the task and for a significant effect of task within levels of site. For all tests, we used the Kenward-Roger method to approximate the denominator degrees of freedom for the F-statistic and considered a significance level of 0.05. All analyses were carried out in R version 4.2.2.

## Data availability

Though the data are not as yet published publicly, the authors will entertain requests for collaboration and data sharing especially if not part of our planned next stage analyses.

## Results

### Mixed models

Since there was variability across patients in the time between baseline symptom assessment and start of treatment that could have affected the trajectory of symptom change across time, the number of days between baseline assessment and treatment initiation was included in each model as a covariate of no interest. Table 1 displays results for main effects of Site (fMRI-guided vs. 6cm scalp target), Task condition (N-back cognitive task vs. nature Video), and their interaction on overall depression, overall PTSD, and PTSD symptom subscales obtained from the linear mixed effect models described above.

**Table 1.** (a) Results from longitudinal models of change from baseline in PHQ, PCL, and PCL subscales without Site-by-Task interaction. For each outcome, change from baseline was modeled using a linear mixed model with subject random intercept and fixed effects for baseline score, time in days from baseline assessment to first treatment visit, time in days since first treatment visit, site main effect (fMRI-guided vs scalp-defined target), and task main effect (N-back vs Video). **(b)** Results from longitudinal model of change from baseline in PCL avoidance subscale with Site-by-Task interaction. Change from baseline was modeled using a linear mixed model with subject random intercept and fixed effects for baseline score, time in days from baseline score collection to first treatment visit, time in days since first treatment visit, site main effect (fMRI-guided vs scalp target), task main effect (N-back vs Video), and site-by-task interaction. **ndf:** numerator degrees of freedom. **ddf:** denominator degrees of freedom. Site was coded as **0=Scalp** and **1=fMRI-guided**. Task was coded as **0=N-back** and **1=Video**.

| (a) Models without Site-by-Task interaction |  |  |  |  |  |  |
| --- | --- | --- | --- | --- | --- | --- |
| Scale | Effect | Estimate | F Stat | ndf | ddf | p-value |
| PHQ | Site | -2.53 | 5.93 | 1 | 43.92 | **0.019** |
|  | Task | 0.42 | 0.16 | 1 | 43.65 | 0.688 |
| PCL | Site | -2.33 | 0.4 | 1 | 39.83 | 0.530 |
|  | Task | 2.12 | 0.33 | 1 | 39.80 | 0.571 |
| PCL: Reexperiencing | Site | -0.04 | 0.00 | 1 | 40.68 | 0.965 |
|  | Task | 1.03 | 1.23 | 1 | 40.64 | 0.274 |
| PCL: Negative Alterations | Site | -0.25 | 0.03 | 1 | 41.94 | 0.875 |
|  | Task | 0.74 | 0.21 | 1 | 41.65 | 0.647 |
| PCL: Hyperarousal | Site | -2.13 | 5.08 | 1 | 40.78 | **0.030** |
|  | Task | 0.23 | 0.06 | 1 | 40.87 | 0.810 |
| (b) Model with Site-by-Task interaction |  |  |  |  |  |  |
| Scale | Effect | Estimate | F Stat | ndf | ddf | p-value |
| PCL: Avoidance | Site-by-Task Interaction | -3.07 | 6.17 | 1 | 40.44 | **0.017** |

### Models for PHQ9

The site-by-task interaction was not significant (*F*(1, 42.33)=2.33; *P*=0.134), so it was dropped from the model. Based on the model with main effects of site and task, the fMRI-guided site was estimated to result in 2.5 point greater reduction in baseline PHQ9 score than the 6cm scalp site, and this effect was significant (*F*(1, 43.92)=5.93; *P*=0.019). The expected difference in reduction from baseline between the cognitive task and video task conditions was only 0.42 points on the PHQ9 score scale, and this effect was not significant (*F*(1, 43.65)=0.16; *P*=0.688). Figure 1 displays marginal effects of the site and task conditions, derived by averaging over all other factors and variables in the mixed model, along with 95% confidence intervals. The superiority of the fMRI-guided site on PHQ9 is reflected by the top interval in Figure 1 which appears in the negative region and does not include zero.

**Figure 1.**
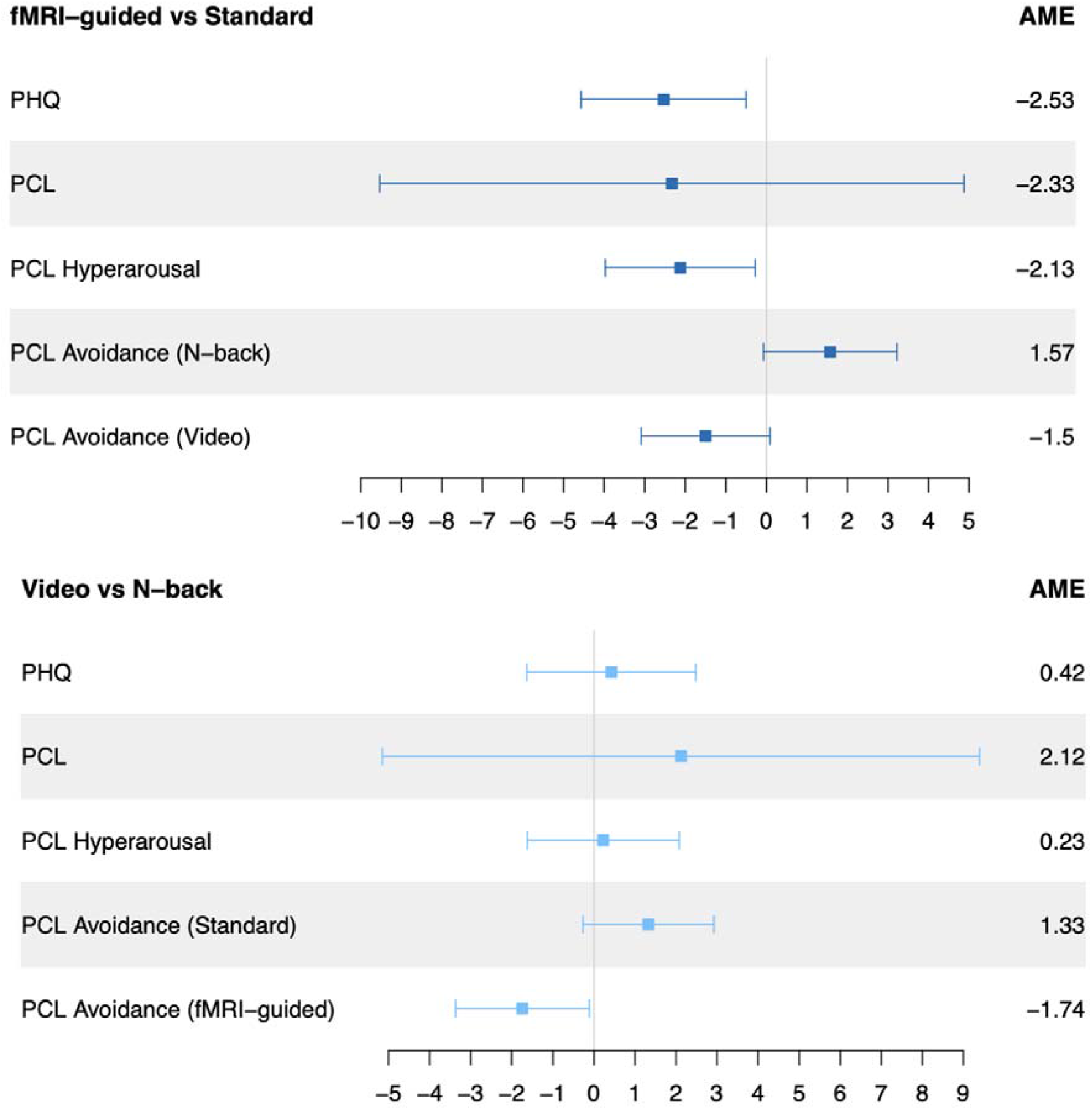
Forest plots of the average marginal effect (AME) and corresponding 95% confidence interval comparing stimulation sites (fMRI-guided vs. scalp target) and concurrent tasks (Video vs. N-back cognitive task) with respect to 4 outcomes: depression (PHQ), PTSD overall (PCL) and the specific subscales of Hyperarousal and Avoidance derived from the PCL. Because of the significant site-by-task interaction, PCL Avoidance appears twice in each panel, i.e., separate AMEs are plotted within subgroups defined by the other factor. In the top panel, negative values indicate superiority of the fMRI-guided site. In the bottom panel, negative values indicate superiority of the video task.

### Models for PCL-5

The site-by-task interaction was not significant (*F*(1, 39.48)=3.10; *P*=0.086), so it was dropped from the model. Based on the model with main effects of site and task, the fMRI-guided site was estimated to result in a 2.3 point greater reduction in baseline PCL-5 score than the scalp-defined site, but this effect was not significant (*F*(1, 39.83)=0.40; *P*=0.530). The task condition was estimated to have a 2.1 point greater reduction in PCL-5 score from baseline than the video condition, but this effect was not significant (*F*(1, 39.80)=0.33; *P*=0.571).

### Models for PCL Sub-scores

We did not find evidence of site or task condition effects on the Re-experiencing or Negative Alterations sub-scores of the PCL-5. For Avoidance, the site-by-task interaction was significant (*F*(1, 40.44)=6.17; *P*=0.017). The estimated coefficient on the interaction term was -3.1, indicating a 3.1 point greater expected reduction from baseline in Avoidance when fMRI-guided was given with the video task compared to the scalp target with cognitive task. In follow-up analyses within levels of site and task condition, we found a significant effect of task within the group who received fMRI-guided stimulation (*F*(1, 18.12)=5.34; *P*=0.033). For that group, the effect estimate corresponded to an expected 2.1 point greater reduction from baseline in Avoidance when patients were exposed to the video task condition.

The site-by-task interaction was not significant in the model for Hyperarousal ((*F*(1, 39.70)=1.71; *P*=0.198), so it was dropped from the model. Based on the model with main effects of site and task, the fMRI-guided site was estimated to result in a 2.1 point greater reduction in baseline Hyperarousal score than the scalp-defined site, and this effect was significant (*F*(1, 40.78)=5.08; *P*=0.030; see Table 1(a) and top panel of Figure 1). The cognitive task condition was estimated to have a 0.23 point greater reduction in Hyperarousal score from baseline than the video condition, but this effect was not significant (*F*(1, 40.87)=0.06; *P*=0.810).

### Outcome Distributions by Time Point

To visualize differences in treatment response over time, Figure 2 displays boxplots of the distribution of complete-case data at each treatment timepoint and the 1^st^ follow-up visit (‘1wk Post’) by target (fMRI-guided vs scalp-guided) for PHQ9, PCL, and the PCL Hyperarousal subscale. These plots do not directly correspond to the average trends captured by the mixed effects models. However, Figure 2 shows greater improvement in median change from baseline over time for the fMRI-guided site compared to the scalp target on PHQ9 and PCL Hyperarousal, corresponding to the two outcomes for which site had a significant main effect in our models.

**Figure 2.**
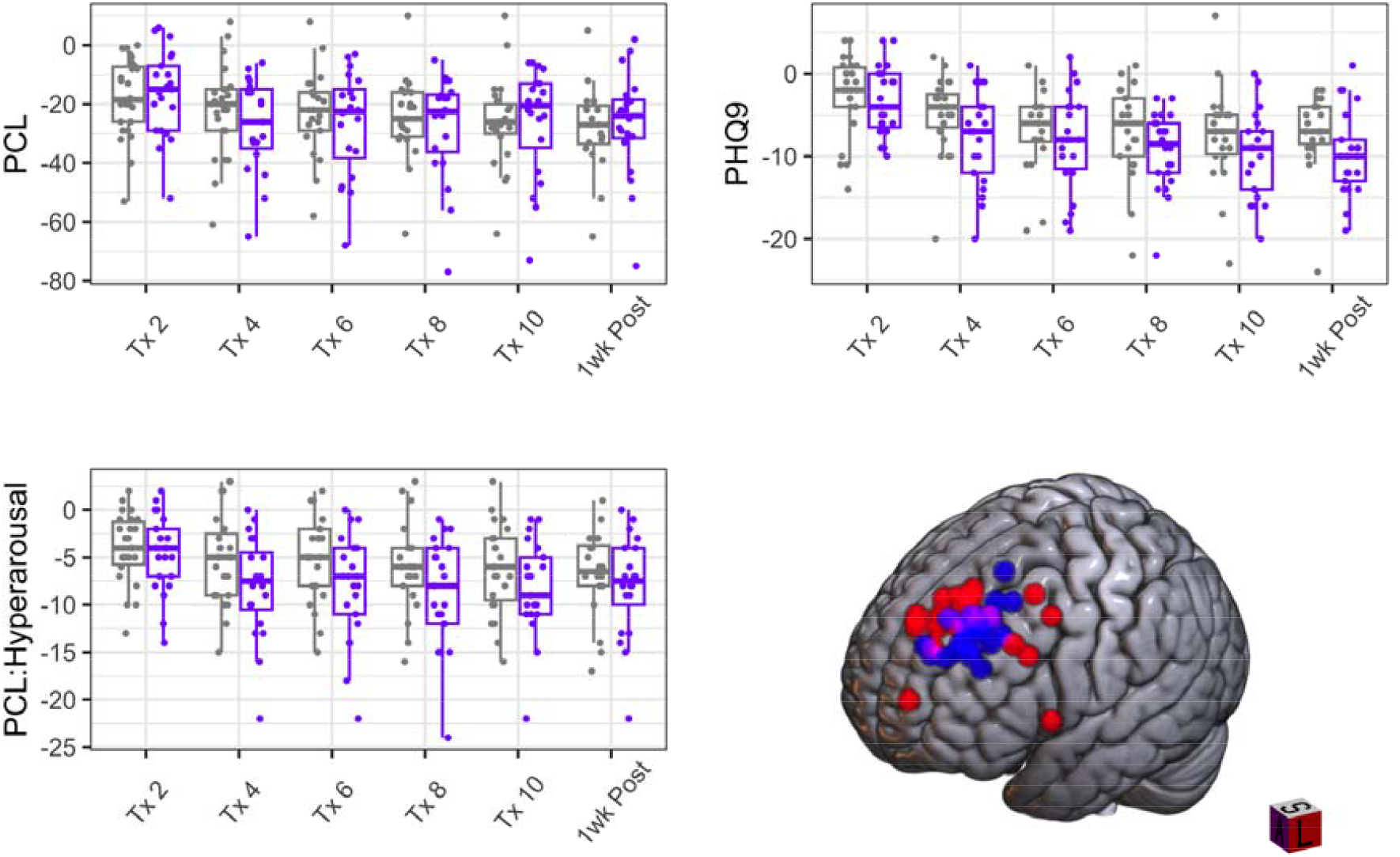
Distributions of change from baseline in PHQ9 (Depression), PCL (PTSD), and PCL Hyperarousal subscale over time by stimulation site (fMRI-guided: purple; Scalp-defined: grey). ‘Tx’ indicates the treatment session number and ‘1wk Post’ indicates the post-treatment check-in 1 week after the last rTMS session to that target (Period 1). Each boxplot is based on available observations at a given time point (i.e., not accounting for missingness). The brain image shows fMRI-guided (red) and scalp-based (blue) targets across patients. The most anterior and lateral two fMRI sites were less spatially anomalous (closer to DLPFC) in native structural space.

Amongst the 43 patients with a PHQ9 score available immediately post-treatment (i.e., post-treatment, session 10 data; complete case analysis), the group assigned to the fMRI-guided target experienced a median 67% reduction from baseline to post-treatment compared with a 50% median decrease in the scalp target group. Using available PHQ9 measures immediately post-treatment, the responder rate (defined as 50% or more reduction from baseline) was 68% in the fMRI-guided group and 59% in the scalp target group.

Amongst patients assigned to the fMRI-guided site, the video group experienced a median 83% improvement in Avoidance compared with the cognitive task (15% median improvement). The scalp-defined site had highly similar Avoidance improvement for video (50% median improvement) but better improvement amongst patients who received the cognitive task (58% median improvement). For Hyperarousal, the fMRI-guided protocol resulted in a median 64% reduction in symptoms whereas the scalp target reduced median symptoms by 43% amongst complete data immediately post-treatment.

### Follow-up Period

Depression improvement in the fMRI-guided group relative to baseline at 1-month and 6-month post-treatment was 58% [N15 patients reporting] and 70% [N13], respectively. This gradual improvement in symptoms could be the result of patients staying in the study who were improving, biasing the results in favor of better outcomes. Across the 6 mo follow up assessments, 5 patients were lost to follow-up who had reported <50% improvement in depression compared with baseline at their last assessment. By contrast, there were 13 patients with at least one assessment showing <50% improvement who completed all follow-up symptom reports through 6 mo post-treatment. For patients *switched* to the fMRI-guided target during the 2^nd^ phase of treatment (following scalp-targeted rTMS), depression improvement was 56% at 1-month [N18] and 44% after 6 mo [N10] post-treatment, representing a loss of 21% of the clinical effect over the extended follow-up period. The PTSD symptom improvement across both sites at 1-month post-treatment was 68% [N31] and 47% at 6 mo [N21]. Though the site difference for the PCL scale was not significant immediately post treatment, the fMRI-guided benefit at 1-month post-treatment (73% improved [N13] vs. scalp 63% [N18]) became more pronounced at 6 mo post-treatment (69% improved [N11] vs. scalp 42% [N10]). This represents <1% loss of treatment benefit for PTSD symptoms in the fMRI-guided group. The hyperarousal and avoidance subscales both showed strong improvements at follow-up (fMRI-guided hyperarousal 78% improved [N14] and fMRI-guided avoidance 79% improved [N12] at 6 mo). In aggregate, the follow-up period results suggest an extended clinical benefit of fMRI-guided rTMS to treat both depression and PTSD. However, in light of the magnitude of missing data throughout the follow-up phase and the potential for patients to pursue additional treatments in the follow-up which we didn’t track, our follow-up results may reflect unmeasured confounding and selection bias induced by non-random missingness. For both depression and PTSD symptom improvement, the highest rate of responders at the follow-up was for fMRI-guided targeting paired with the relaxation video task condition reaching nearly 75% for each symptom category (Supplementary Figures 3-4).

## Discussion

Although a number of laboratories are pursuing individual neuroimaging-based targets for treating psychiatric disorders,^3,4,21,22^ it has not yet been demonstrated that a head-to-head comparison between image-guided compared with non-image guided targeting demonstrates the predicted increase in clinical effectiveness for the more personalized image-guided rTMS treatment.^1,2^ One might assume the superiority of image-guided rTMS for treating depression given unprecedented clinical outcomes following the recently FDA approved protocol.^3,4^ However, this new rTMS protocol differs from published protocols in terms of a number of procedural elements, timing of sessions, and number of overall rTMS pulses that might partially account for the superior clinical effects beyond the image-based targeting approach. Motivated by this gap in the literature, one goal of the present study was to compare the effectiveness of a scalp-based target to our own novel fMRI-guided rTMS target with respect to symptom improvement over time. We have previously demonstrated the promise of our novel fMRI-guided rTMS target over anticorrelated subgenual anterior cingulate targets to engage the sgACC in patients, as measured by online interleaved TMS/fMRI.^5^ On the clinical side, our symptom reduction for the fMRI-guided protocol was 67% for depression, which falls in the range of the latest Stanford (SNT) protocol (62-86% reduction^3,4^), outstrips the recent fMRI-guided BRIGhTMIND British protocol (37% reduction^2^), and the Canadian THREE-D structural MRI based target (43% reduction^1^). This improvement is also substantially better than their F3 scalp target (35% reduction) as well as the original 5cm target from Penn (18-23% reduction^16^). Our follow up data are comparable to SNT (1-mo 52.5-69% symptom reduction) for both depression (70% improved for fMRI target) and PTSD (69% improved for fMRI target) even comparing our *6 mo* results to the prior study *1 mo* results. It should be noted that the dose for the Stanford (90k), original 5cm (60-90k) and recent BRIGhtMIND protocols (63k), are higher than the THREE-D (12-18k) as well as our protocol (23k per site). Thus, it is possible that our rTMS protocol could yield even stronger clinical effects with additional treatment sessions. In addition to testing higher dose clinical interventions, we are inclined to pursue mechanistic studies of how TMS engages brain circuits and how clinical rTMS modulates communication through these circuits through the use of interleaved TMS/fMRI. For example, we have demonstrated circuit engagement at the individual patient level supporting symptom-specific pathway modulation with TMS^14^ with other groups yielding parallel evidence for using correlative imaging and lesion methods.^23^

Limitations include our mixed PTSD/MDD sample with insufficient numbers of patients with only one diagnosis limiting our ability to make predictions for single diagnosis patient cohorts. Due to small cell sizes per condition with full diagnoses, remission rates here are less informative and so were not calculated. Also, we did not require patients to be treatment resistant, as most prior rTMS treatment studies have, but 32.5% of the sample in the post-treatment analyses met this criterion (non-response to 2+ antidepressant medications). Since the cross-over wash-out period was insufficient to return symptoms to baseline before phase 2, it is possible that carry-over effects influenced the 2^nd^ treatment period clinical outcomes. Future studies could either limit treatment to a single 2-week target or extend the dose in a longer parallel study to test the outcome of separate targets and task conditions.

These findings represent evidence that an fMRI-guided rTMS target is clinically superior to a less individualized scalp measurement-based target for treating depression and PTSD (though PTSD advantages were in long-term follow-up or immediate for a specific subscale). In addition, findings suggest that there may be an interaction between brain state (assumed with psychological task manipulation) at the treatment sessions and clinical improvement for some symptoms of PTSD. Clinical benefits of this novel fMRI target were durable across 1- and 6-months post-treatment even without additional rTMS treatment. Precision, personalized image-guided brain stimulation targeting is making strong headway into clinical rTMS practice in psychiatry.^3,4^ Additional work exploring other targets and indications is underway and showing promising preliminary results.^23-25^

## Funding

CureAccelerator Repurposing Award from Cures Within Reach and the Kahlert Foundation (DJO).

## Competing interests

The authors report no competing interests.

## Supplementary material

Supplementary material is available online.

## Notes

### Competing Interest Statement

The authors have declared no competing interest.

### Clinical Trial

NCT03114891

### Author Declarations

The Institutional Review Board of the University of Pennsylvania Perelman School of Medicine gave ethical approval for this work

