## Supplemental Materials for "Clinical Response to fMRI-guided Compared to Non-Image Guided rTMS in Depression and PTSD: A Randomized Trial"

### Introduction

### Materials and methods

**Patients**

Participants were excluded if they had MRI contraindication (implanted devices not MRI safe, claustrophobia, etc.), TMS counterindications (seizure disorder, CNS active disorder, medication use that substantially reduces seizure threshold to TMS), pregnancy risk, other neurological disorders including multiple sclerosis, encephalopathy, or brain tumors. They were also excluded if they met criteria for a current moderate or severe alcohol or substance abuse disorder, bipolar disorder, schizophrenia or other psychotic disorder, pregnancy, or previous unsuccessful treatment with a full trial of TMS, DBS, or Electroconvulsive therapy (ECT). Two patients did not provide adequate data to be included in the primary analysis models. See **Supplementary Figure 1** for a CONSORT diagram detailing study stages and patient inclusion.

**Supplementary Figure 1.** CONSORT diagram detailing number of patients assessed, enrolled, randomized to treatment, and completed for each study arm.

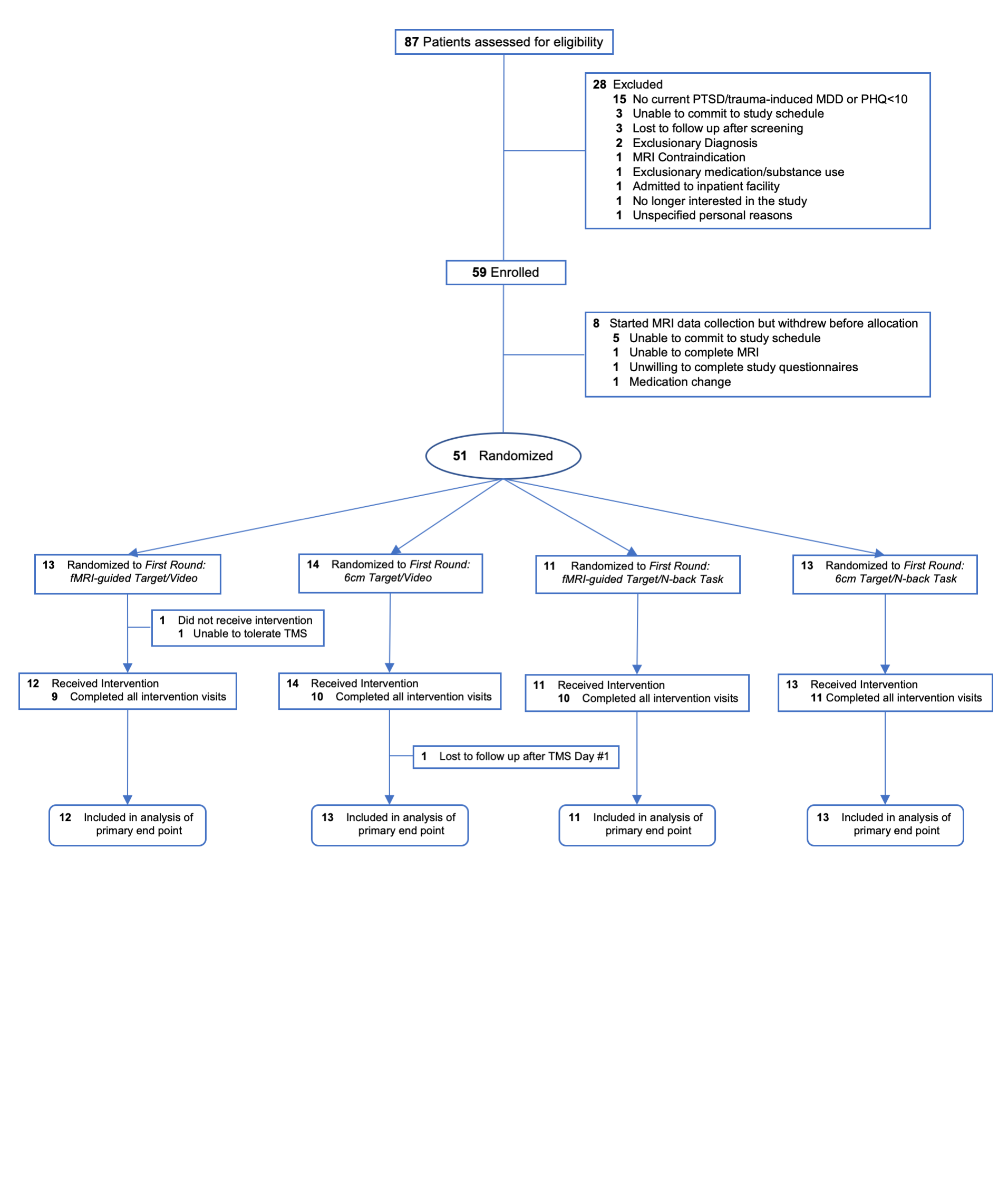

Demographics, baseline symptom levels, missed sessions, and principal diagnoses are split according to the comparison of fMRI-guided or scalp (standard) target (**Supplementary Table 1**) then in a 2^nd^ table additionally divided according to which task was utilized during rTMS sessions (**Supplementary Table 2**) for the first period treatment protocol.

| **Supplementary Table 1** | | | | | |
| --- | --- | --- | --- | --- | --- |
| **Characteristics** | **All (n=49)** |  | **fMRI-guided (n=23)** |  | **Standard (n=26)** |
|  | **Mean (SD)** |  | **Mean (SD)** |  | **Mean (SD)** |
| Age | 34.04 (11.18) |  | 32.39 (11.70) |  | 35.50 (10.71) |
| Total Traumas | 3.94 (2.24) |  | 3.65 (2.35) |  | 4.19 (2.15) |
| Baseline PHQ | 16.76 (4.36) |  | 16.74 (4.55) |  | 16.77 (4.27) |
| Baseline PCL | 50.17 (13.39) |  | 49.33 (14.76) |  | 50.85 (12.44) |
| Number of Missed Treatments | 4.67 (5.44) |  | 4.09 (5.11) |  | 5.19 (5.77) |
|  | **Percentage** |  | **Percentage** |  | **Percentage** |
| Sex Assigned at Birth |  |  |  |  |  |
| Male | 35.4 |  | 30.4 |  | 40.0 |
| Female | 64.6 |  | 69.6 |  | 60.0 |
| Current Gender |  |  |  |  |  |
| Male | 34.7 |  | 30.4 |  | 38.5 |
| Female | 59.2 |  | 60.9 |  | 57.7 |
| Do not identify as   Male, Female, or Transgender | 6.1 |  | 8.7 |  | 3.8 |
| Highest Education |  |  |  |  |  |
| GED or equivalent | 2.0 |  | 0.0 |  | 3.8 |
| High School Graduate | 4.1 |  | 8.7 |  | 0.0 |
| Some College | 30.6 |  | 21.7 |  | 38.5 |
| Associate's Degree | 4.1 |  | 0.0 |  | 7.7 |
| Bachelor's Degree | 42.9 |  | 47.8 |  | 38.5 |
| Master's Degree | 12.2 |  | 17.4 |  | 7.7 |
| Professional Degree | 2.0 |  | 0.0 |  | 3.8 |
| Doctoral Degree | 2.0 |  | 4.3 |  | 0.0 |
| Principal Diagnosis |  |  |  |  |  |
| MDD Only | 26.5 |  | 34.8 |  | 19.2 |
| PTSD Only | 69.4 |  | 65.2 |  | 73.1 |
| Both MDD and PTSD | 4.1 |  | 0.0 |  | 7.7 |

**Supplementary Table 1.** Patient descriptions for the full (‘All’) sample then divided according to whether their first treatment target was fMRI-guided or the ‘Standard’ (6 cm scalp) target. Mean and Standard Deviation (SD) are provided, as appropriate.

**Supplementary Table 2.** Patient descriptions for the full (‘All’) sample then divided according to whether their first treatment target was fMRI-guided or the ‘Standard’ (6 cm scalp) target as well as according to the Task they were asked to engage in during rTMS sessions (‘N-Back’ working memory [cognitive] task or ‘Video’ natural scene videos). Mean and Standard Deviation (SD) are provided, as appropriate.

| **Supplementary Table 2** | | | | | | | | | |
| --- | --- | --- | --- | --- | --- | --- | --- | --- | --- |
| **Characteristics** | **All  (n=49)** |  | **fMRI-guided + N-Back (n=11)** |  | **fMRI-guided + Video (n=12)** |  | **Standard + N-Back (n=13)** |  | **Standard + Video (n=13)** |
|  | **Mean (SD)** |  | **Mean (SD)** |  | **Mean (SD)** |  | **Mean (SD)** |  | **Mean (SD)** |
| Age | 34.04 (11.18) |  | 29.27 (8.45) |  | 35.25 (13.79) |  | 36.00 (10.57) |  | 35.00 (11.25) |
| Total Traumas | 3.94 (2.24) |  | 4.36 (2.38) |  | 3.00 (2.22) |  | 3.69 (2.14) |  | 4.69 (2.14) |
| Baseline PHQ | 16.76 (4.36) |  | 18.27 (3.85) |  | 15.33 (4.85) |  | 17.31 (3.45) |  | 16.23 (5.05) |
| Baseline PCL | 50.17 (13.39) |  | 56.09 (14.32) |  | 41.90 (11.79) |  | 49.08 (13.39) |  | 52.62 (11.67) |
| Number of Missed Treatments | 4.67 (5.44) |  | 4.27 (4.96) |  | 3.92 (5.45) |  | 4.69 (5.07) |  | 5.69 (6.56) |
|  | **Percentage** |  | **Percentage** |  | **Percentage** |  | **Percentage** |  | **Percentage** |
| Sex Assigned at Birth |  |  |  |  |  |  |  |  |  |
| Male | 35.4 |  | 27.3 |  | 33.3 |  | 38.5 |  | 41.7 |
| Female | 64.6 |  | 72.7 |  | 66.7 |  | 61.5 |  | 58.3 |
| Current Gender |  |  |  |  |  |  |  |  |  |
| Male | 34.7 |  | 27.3 |  | 33.3 |  | 30.8 |  | 46.2 |
| Female | 59.2 |  | 54.5 |  | 66.7 |  | 61.5 |  | 53.8 |
| Do not identify as Male, Female, or Transgender | 6.1 |  | 18.2 |  | 0.0 |  | 7.7 |  | 0.0 |
| Highest Education |  |  |  |  |  |  |  |  |  |
| GED or equivalent | 2.0 |  | 0.0 |  | 0.0 |  | 7.7 |  | 0.0 |
| High School Graduate | 4.1 |  | 9.1 |  | 8.3 |  | 0.0 |  | 0.0 |
| Some College | 30.6 |  | 36.4 |  | 8.3 |  | 30.8 |  | 46.2 |
| Associate's Degree | 4.1 |  | 0.0 |  | 0.0 |  | 0.0 |  | 15.4 |
| Bachelor's Degree | 42.9 |  | 45.5 |  | 50.0 |  | 46.2 |  | 30.8 |
| Master's Degree | 12.2 |  | 9.1 |  | 25.0 |  | 7.7 |  | 7.7 |
| Professional Degree | 2.0 |  | 0.0 |  | 0.0 |  | 7.7 |  | 0.0 |
| Doctoral Degree | 2.0 |  | 0.0 |  | 8.3 |  | 0.0 |  | 0.0 |
| Principal Diagnosis |  |  |  |  |  |  |  |  |  |
| MDD Only | 26.5 |  | 45.5 |  | 25.0 |  | 15.4 |  | 23.1 |
| PTSD Only | 69.4 |  | 54.5 |  | 75.0 |  | 84.6 |  | 61.5 |
| Both MDD and PTSD | 4.1 |  | 0.0 |  | 0.0 |  | 0.0 |  | 15.4 |

Antidepressant Treatment Resistance: 19 patients in the analysis sample reported having taken 2+ antidepressants in the past with inadequate relief from symptoms; the median number of medications taken in the past was 3, with a range of 2-15+ among these patients; 3 additional patients reported having benefited from antidepressants in the past but were not currently taking them for a variety of reasons (personal preference, insurance/financial, etc.).

**MRI acquisition and processing**

For each participant, a resting state fMRI scan was acquired on a Siemens Prisma 3 Tesla whole-body MRI with a 64-channel head coil (Erlangen, German) (phase encoding direction: anterior to posterior, TR=720ms, TE=37ms, FA=52°, FOV=208mm, 2×2×2mm voxels, 72 interleaved axial slices with no gap, 600 measurements). During the functional scan, participants were instructed to keep their eyes open and remain as still as possible. Structural data consisted of a high-resolution multi-echo T1-weighted MPR image (TR=2400ms, TI=1060ms, TE=2.24ms, FA=8°, 0.8×0.8×0.8mm voxels, FOV=256mm, PAT mode GRAPPA, 208 slices) were also acquired. For resting fMRI data processing, the first 10 volumes were discarded for T1 equilibrium, and then automated removal of motion artifact designed to maintain fMRI autocorrelation structure using ICA-AROMA^1^ was applied. Nuisance regression, residualizing for white matter and CSF signal, was implemented in FSL 5.08 (FMRIB Oxford, UK), followed by band-pass filtering 0.008–0.1 Hz and 6 mm kernel FWHM smoothing to fMRI data. Boundary-based registration following FSL FAST tissue segmentation used six DOF to coregister T1 to functional scans and FSL FNIRT default settings were used for nonlinear warps of fMRI data to 2 mm MNI152 template space. The inverse of this process moved seed regions to native fMRI space, and functional connectivity for each seed was calculated in native space, resulting in Pearson correlation maps that were transformed to z scores using the Fisher’s r-to-z equation. Inverting the T1 to functional transform placed the fMRI connectivity maps in native T1space for neuronavigation that were visually verified.

**TMS treatment, additional details**

The motor threshold was determined with the TMS coil positioned at the primary motor cortex (M1) with the coil handle facing backward. The mapping of M1 was guided by the hand knob landmark from the structural image, and the motor threshold was identified by the muscle twitches (5/10 trials) of the FDI (first dorsal interosseuous) or APB (abductor pollicis brevis) of participant’s dominant/right hand (whichever most clearly responded) evoked by TMS.

Each round of treatment was administered over ten treatment visits (generally Monday through Friday). Visits were scheduled as close to one another as possible (consecutive days), yet some visits were a few days apart from each other due to weekends and patient availability.

**Brain state manipulations**

Arousal (in the context of reward) has been shown to be influenced by the sgACC in monkeys^2^ and was shown to influence mood effects of electrical stimulation to sgACC in an N-of-1 patient study.^3^ It is been long known that psychological manipulations influence TMS-induced motor responses^4^ and more recently have been applied in rTMS treatments of MDD^5^, PTSD^6^, OCD^7^ and smoking cessation.^8^ However, clinical studies have shown mixed results across PTSD^6^ and MDD^5^ with symptom provocations combined with rTMS. We expanded the repertoire of these initial context manipulations here to test interactions between brain stimulation target, effort/arousal and symptom improvement for depression and PTSD.

Each treatment session consisted of two rounds (800 triplets/round), and putative brain states were induced by asking participants to either complete the fractal n-back task^9^ or view a nature video between the two rounds (<https://www.youtube.com/watch?v=nxJgR33CVs4> + <https://www.youtube.com/watch?v=aS-LUW5Jim0>). The version of the n-back task utilized a constant display of neutral fractal images and required participants to provide a response when a new image matched the image that was shown *n* trials before the current trial (n = 0, 1, 2). There were ~10 minutes between the two rounds of TMS. Since the fractal n-back task requires participants to keep track of the order of previous images while attending to new images, it was intended to engage cognitive control circuits and reflect effortful task engagement. The video included peaceful clips of nature scenes, and served to induce relaxed, low-arousal brain states. Each participant experienced both induced brain states with the sequence of states randomly assigned to participants as part of the crossover design (task in period 1 and video in period 2 or vice versa). In each period of the crossover design, the same brain state was induced during every treatment visit for each round (10 visits).

### Results

**Evidence of Carryover Effects**

The mean PHQ9 prior to period 2 (i.e., at the second check in visit) was significantly lower than the mean PHQ9 prior to period 1 (*t*(39)=-9.45; *P*<0.001). The mean PCL prior to period 2 was significantly lower than the mean PCL prior to period 1 (*t*(38)=-10.45; *P*<0.001).

**Follow-up Data**

Not all patients were reachable across the 6-month follow up period; at 1-month post-treatment, we were able to obtain 33 PHQ9 scores and 31 PCL-5 scores. By 6-month follow-up, we collected 23 PHQ9 and 21 PCL-5 scores.

**Response (≥50% improvements)**

Response proportion trajectories are plotted for the purpose of suggesting priorities to test in replication studies according to stimulation site and concurrent task separately for depression and PTSD symptom changes across the treatment period and into the post-treatment 1-week follow-up period. The results support use of the fMRI target for depression (Supplementary Figure 2A) and warn against combining the scalp target with the nature video (Supplementary Figure 2B).

**Supplementary Figure 2A/B.** Depression (‘PHQ9’) proportion responder (≥50% improvement in symptoms) numbers (with 95% pointwise confidence intervals) steadily increased over time (‘Tx’ Treatment day) into the post-treatment 1-week follow-up (‘Post Tx’) period that stayed steady for the fMRI-guided target but declined for the scalp (‘Standard’) target at follow-up (A). Though the fMRI-guided target paired with the Video condition had the ultimate best depression outcomes numerically at follow-up (B), the Standard Target combined with the Video condition resulted in an especially low response proportion and eventual drop in responder status.

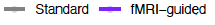

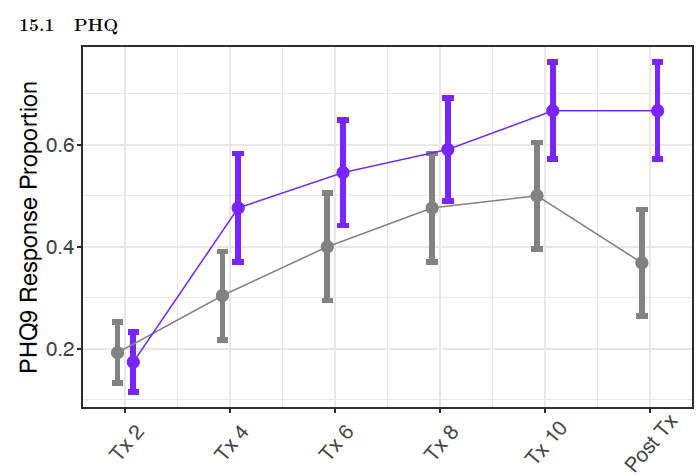

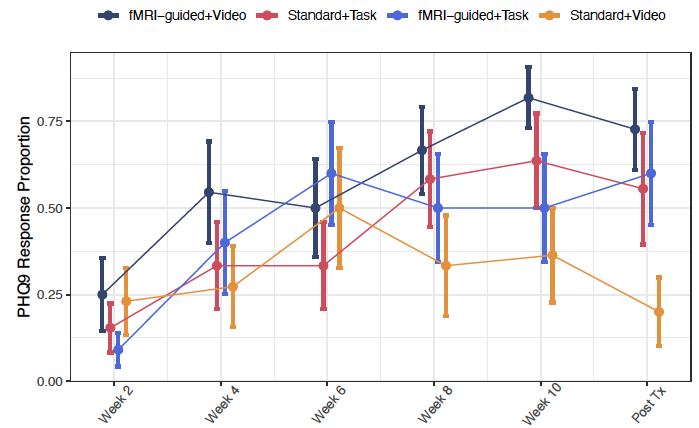

For PTSD ('PCL’) symptom improvement, the results suggesting fMRI-guided target benefits emerging in the post-treatment follow-up (Supplementary Figure 3A) seemed to have been driven especially by the fMRI-guided target paired with the Video condition maintaining the clinical benefit at 1-month post-treatment (Supplementary Figure 3B).

**Supplementary Figure 3A/B.** PTSD (‘PCL’) proportion responder (≥50% improvement in symptoms) numbers (with 95% pointwise confidence intervals) increased over time (‘Tx’ Treatment day) into the post-treatment 1-week follow-up (‘Post Tx’) period (A) that was maintained especially for the fMRI-guided target paired with the Video condition (B).

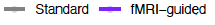

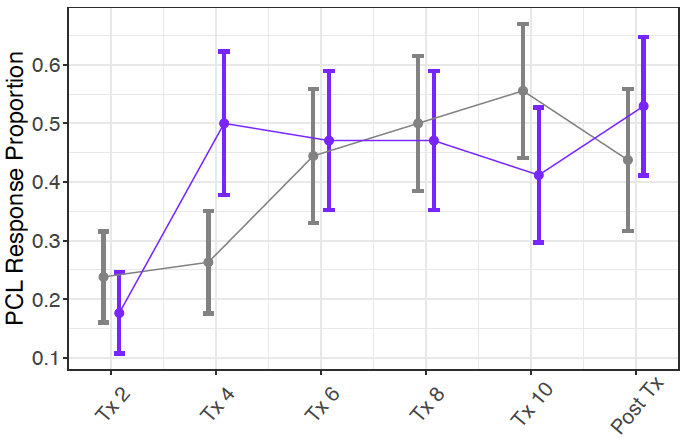

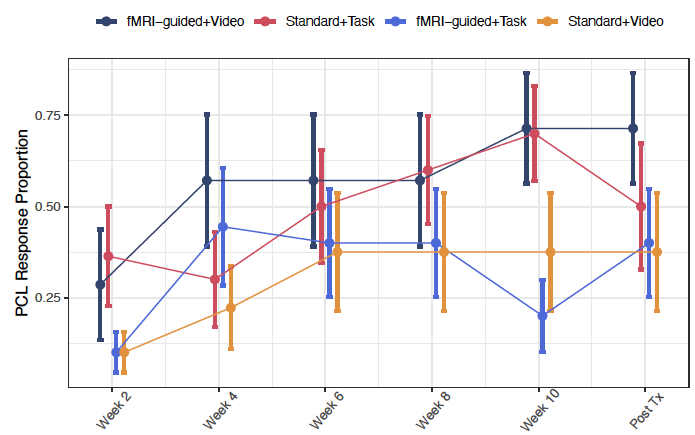

1.	Pruim RH, Mennes M, Buitelaar JK, Beckmann CF. Evaluation of ICA-AROMA and alternative strategies for motion artifact removal in resting state fMRI. *NeuroImage.* 2015;112:278-287.

2.	Alexander L, Gaskin PL, Sawiak SJ, et al. Fractionating blunted reward processing characteristic of anhedonia by over-activating primate subgenual anterior cingulate cortex. *Neuron.* 2019;101(2):307-320. e306.

3.	Scangos KW, Makhoul GS, Sugrue LP, Chang EF, Krystal AD. State-dependent responses to intracranial brain stimulation in a patient with depression. *Nature Medicine.*1-3.
